# A decision-only model as a low-cost judge of medical AI answers

**DOI:** 10.64898/2026.10.30.26366939

**Authors:** Mahmud Omar, Benjamin S. Glicksberg, Ankit Sakhuja, Yiftach Barash, Alexander W. Charney, Eyal Klang, Girish N. Nadkarni

**Author notes:** Corresponding author - Mahmud Omar, MD.

## Abstract

**Background:** Testing and deploying medical AI systems at scale already results in more outputs than clinicians can review, and the use of large language models (LLMs) as judges is growing rapidly. Decision-only models, cheap and fast models that return structured probabilities rather than text, offer a potentially scalable option for that goal.

**Methods:** In an OSF pre-registered evaluation, we tested TypeSafe Jev 1.13 as a judge on tasks from five validated clinical benchmarks: rubric grading (HealthBench), planted clinical errors (When Rubrics Fail), risk in AI-written clinical documents (MedVAL), counseling ratings and failures (CounselBench) and physicians’ preferences between answers (Real-POCQi). Comparators were expert raters’ agreement with each other, OpenAI Decisions, GPT-6 Luna, a proposed hybrid team combining decision models and a cheap LLM and, as frontier comparators, GPT-6.1 Sol and Claude Opus 5.5. The main outcome was plain agreement with the expert label.

**Results:** The test comprised 25,091 units and 213,913 model calls. Jev’s agreement with expert labels averaged 64.7% over six tasks. On 3,614 matched units it averaged 66.1%, against 65.0% for Opus and 60.7% for Sol, and it was non-inferior to both on four of six tasks. Jev judged the whole pool in 17 minutes for US$1.46; per judgment, it cost 1/34 of Sol’s price and 1/152 of Opus’s. A preset cutoff held its false-alarm rate on fresh planted-error answers, and in simulation, reviewing Jev’s least-sure answers first found 1.4 times the errors of a random audit. Still, omissions were hard for both: at the same false-alarm rate, Jev missed 55.6% of planted omissions and Opus 48.1%, and Opus caught 8.8 points more planted errors overall.

**Conclusions:** Across more than 200,000 model calls, a decision-only model was non-inferior to two frontier judges on most tasks at a small fraction of their cost and time, while reporting how sure it was of every call. With local cutoffs and expert oversight of omissions and clinical documents, judging every output of a deployed medical AI system becomes more affordable.

## Introduction

Medical artificial intelligence (AI) now writes answers faster than anyone can check them. In real-world use, health systems are already deploying large language models (LLMs) and AI agents to draft replies to patient messages, write clinical notes from recorded visits and answer clinicians’ questions at the point of care (1–6). As these systems gain capability and autonomy, their output is growing in scale: one health system logged 2.5 million uses of an AI scribe in its first year (3,4). Each of these outputs needs evaluation and monitoring. Not only is this scale impractical for continuous manual evaluation, but data shows that clinicians who review AI drafts still miss many of the errors in them (7). Thus, more scalable and efficient judge models, and frameworks for collaborative work with physicians, are needed.

LLMs used as automated judges are the main answer so far, and they already run at scale (8–10). HealthBench, a widely used benchmark of health conversations, grades each answer with an LLM that checks it against physician-written criteria, called rubric items; on some themes that grader agrees with physicians about as often as physicians agree with each other (11). Yet the compute, time and cost of judging grow with every answer judged. The strongest current judges, mostly frontier LLMs, might be costly and slow. In addition, LLM judges show tendencies to return a different verdict when asked again and to favor longer answers, the answer shown in a particular position and answers from their own model family (12–15).

Decision-only models are a recently introduced and much cheaper interface. Rather than writing text, they return a probability for each option the user specifies (16,17). Jev, from TypeSafe, is priced at US$0.042 per million input tokens, with no charge for output, and answers in well under a second (16). For reference, Claude Opus 5.5 is priced at US$4 per million input tokens and US$20 per million output tokens (18,19), so a judgment can cost a hundredth as much or less. Beyond cost, asked whether an answer contains a clinically consequential error, a decision model returns the probability that it does. That probability makes the judge’s uncertainty visible, which a bare written verdict does not (20,21), and it could enable a health system or a research team to set a cutoff at the false-alarm rate it can afford and send clinicians first to the answers the judge is least sure about (22–24). Early studies, including our own, have tested decision models on clinical decisions, medical questions and general judging tasks, with promising but mixed results (25–27). Whether they can judge open-text medical answers about as well as clinicians and frontier judges has not been tested.

We tested this in a pre-registered evaluation of TypeSafe Jev 1.13 and compared its performance and agreement with experts with those of current frontier LLMs from different providers. We also propose a practical workflow, Dial-and-Review, to decide which answers Jev flags and which ones clinicians check first.

## Methods

### Study design and pre-registration

We split each benchmark by underlying question, so that no question appeared in more than one part: 15% for development, 20% of the rest for validation, and the remainder, about two-thirds of each benchmark, as the held-out test pool. Over eight development rounds and about 60,000 model calls, we fixed the judges’ questions and prompts, built and tested cheap model teams, and set the cutoffs described below (**Supplementary Methods S1**). We pre-registered the test on the Open Science Framework (OSF) on 20 October 2026, fixing the hypotheses, the analysis plan and the cutoffs (**Supplementary Methods S3**).

### Benchmarks

We used five published benchmarks with expert labels, each designed or used to test automated judges, giving six judging tasks (**Figure 1A**; **Supplementary Table S1** and **Supplementary Figure S1**) (5,11,28–30). Each asks a question a clinician would ask of an AI answer:

**Figure 1.**
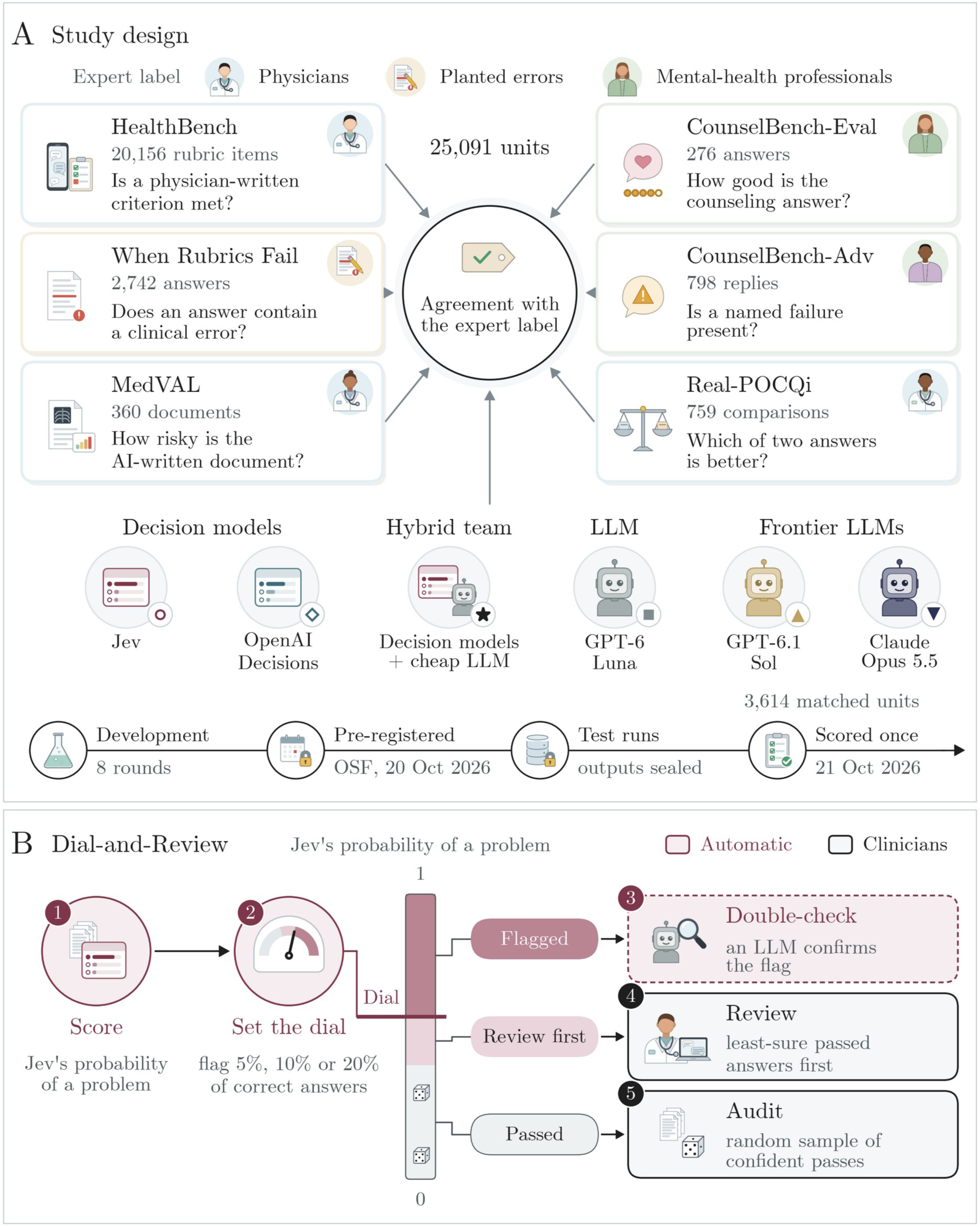
Study design and Dial-and-Review. A, The six tasks, the judges and the timeline. B, The pre-registered workflow for using Jev, and where each step acts on Jev’s probability (schematic); the dashed step is optional.

HealthBench meta-evaluation: does an answer in a health conversation meet a physician-written criterion, such as advising emergency care? Physicians graded each criterion (20,156 rubric items from 2,504 conversations). When Rubrics Fail: does an answer contain a clinically consequential error? Each answer appears in its original form and with one error planted by an LLM, such as a wrong dose, a missed contraindication or an omitted safety step (2,742 answers from 641 questions).

MedVAL: how risky is an AI-written clinical document, such as a radiology impression or a note written from a recorded visit, against its source? Physicians graded risk on four levels (360 documents). CounselBench-Eval: how good is an answer to a mental-health question, on scales such as empathy and factual correctness? Five mental-health professionals rated each answer (276 answers, 1,380 ratings). CounselBench-Adv: does a reply show a named failure, such as inappropriate medication advice? Mental-health professionals labeled each reply (798 replies). Real-POCQi: which of two AI answers to a real clinician question is better, on axes such as accuracy and clinical utility? Physicians chose on each axis (759 comparisons, 3,795 axis ratings).

The test pool held 25,091 units, and identical inputs were judged once. As their authors request, we never display items from HealthBench Professional (31), which enter through When Rubrics Fail, and we did not modify CounselBench items. All benchmarks are public, contain no identifiable patient data and were used within their licenses; no participants were recruited, and institutional review board approval was not required.

### Judges

All judges received the same case text. Jev (TypeSafe Jev 1.13, snapshot 1.13-20260917) and OpenAI Decisions, which OpenAI lists as running on GPT-6 Luna, chose among fixed answer options (yes or no, a rating scale, or a preference between two answers) and returned a probability for each (16,17). For When Rubrics Fail, for example, the question was “Does this answer contain a clinically consequential error, such as an incorrect number, dose, timing threshold, diagnosis, contraindication or instruction, that could change diagnosis or management or cause harm?” The questions used each benchmark’s own wording where it existed and were fixed in the first development round (**Supplementary Table S2**). Jev’s default reading was its most probable option, so it flagged a problem at a probability of 0.5 or more.

GPT-6 Luna, GPT-6.1 Sol and Claude Opus 5.5 received each benchmark’s published judge prompt where one existed (HealthBench, MedVAL and CounselBench-Eval), and otherwise the same question as text, and returned a written verdict that we read with fixed rules. Luna answered three times at low reasoning effort and the majority counted; Sol and Opus answered once at their default settings. Unreadable answers and refusals counted as missing (**Supplementary Table S20**). Frontier models were tested on a matched subset of 3,614 units, drawn at random before any frontier call. Head-to-head comparisons used these units for every judge.

We also tested the Hybrid team, the best cheap team from development, which pairs the decision models with a cheap LLM. It keeps Jev’s verdict and adds a second opinion only where development showed a gain. For instance, on When Rubrics Fail a flag from Jev stands only if deleting one passage of the answer lowers Jev’s error probability by at least 0.10 and Luna confirms that the passage is wrong (the double-check); on MedVAL the team averages Jev’s and Decisions’ probabilities, and on Real-POCQi it averages Jev’s probabilities over both answer orders (**Supplementary Table S2**). Five further team designs from development were tested as pre-registered (**Supplementary Methods S2**).

### Outcomes

The main outcome was plain agreement with the expert label, in percent: agreement with each of the first two physicians on HealthBench, a rating within half a point of the raters’ mean on CounselBench-Eval, the same direction of preference on Real-POCQi, and right calls on the other three tasks (**Supplementary Methods S4**). Where a second expert rating existed (HealthBench, CounselBench-Eval and Real-POCQi), agreement between experts serves as context. Because plain agreement rewards a judge for answering with the most common label, we added after scoring the best constant answer, the single answer that maximizes agreement on each task. The pre-registered comparisons used each benchmark’s own measure (**Table 2**). Secondary outcomes were cost per 1,000 judgments, from recorded tokens at the providers’ prices (16–19), time per call, and consistency on repeated identical requests. We also gave the frontier judges Jev’s exact question and probed every judge for memorized benchmark items (**Supplementary Tables S12 and S13**).

### Dial-and-Review

Dial-and-Review is the pre-registered workflow for using Jev (**Figure 1B**). Jev scores every answer. On the three tasks that ask whether a problem is present (When Rubrics Fail, MedVAL and CounselBench-Adv), an answer is flagged when Jev’s probability of a problem reaches a cutoff, the dial, set on development answers to flag 5%, 10% or 20% of correct answers (the false-alarm rate). To see what a new user would get, we also set the dial on 250 random test answers and tested it on the rest, 1,000 times. The proposed clinician review then goes first to the passed answers closest to the cutoff, at budgets of 1%, 2% and 5% of answers, compared with a random audit of the same size; on the other tasks, review goes to the units where Jev is least sure. For this test, the review process itself was simulated with the benchmark labels (**Supplementary Tables S8 and S9**).

### Pre-registered hypotheses and statistics

We pre-registered four hypotheses: on When Rubrics Fail, the dial would flag within 3 points of each target; at the 10% dial and a 2% budget, review ordered by Jev’s uncertainty would find more errors than a random audit; Jev would be non-inferior to Sol and to Opus on each task in the benchmark’s own measure, within margins fixed in advance (**Table 2**); and the double-check would lower false alarms. Intervals came from a bootstrap that resampled whole questions, conversations, documents or posts 2,000 times. The 14 tests behind the last three hypotheses were adjusted together with the Holm method at a one-sided family-wise level of 0.025, and missing verdicts counted against a hypothesis; the dial hypothesis was judged on the observed share (**Supplementary Methods S3** and **Table S6**). All other comparisons are descriptive, with unadjusted 95% confidence intervals (CIs). Analyses added after scoring, including the best constant answers, the six-task averages, the pooled cost per 1,000 judgments with Sol at batch prices, and the cost and time of judging the whole pool, used the saved outputs without new model calls and are exploratory (**Supplementary Methods S5**).

## Results

### Scale and main results

The test comprised 25,091 units from six tasks and 213,913 model calls, at a recorded cost of US$78 (**Supplementary Table S14**); with development, the study made about 274,000 model calls. Over the six tasks, Jev’s plain agreement with the expert label averaged 64.7% (95% CI 63.4 to 66.0), against 59.5% for the best constant answer, 59.6% for OpenAI Decisions, 62.7% for GPT-6 Luna and 64.1% for the Hybrid team (**Table 1**; **Supplementary Table S3** and **Supplementary Figure S2**). On the 3,614 matched units, Jev averaged 66.1%, against 65.0% for Opus and 60.7% for Sol (**Figure 2**), and in each benchmark’s own measure it was non-inferior to both frontier judges on four of six tasks (**Table 2**). Jev judged all 25,091 units in 17 minutes for US$1.46. Judging the same inputs would have cost an estimated US$55.4 with Sol at batch prices, taking about 4.5 hours, and US$253.4 with Opus.

**Figure 2.**
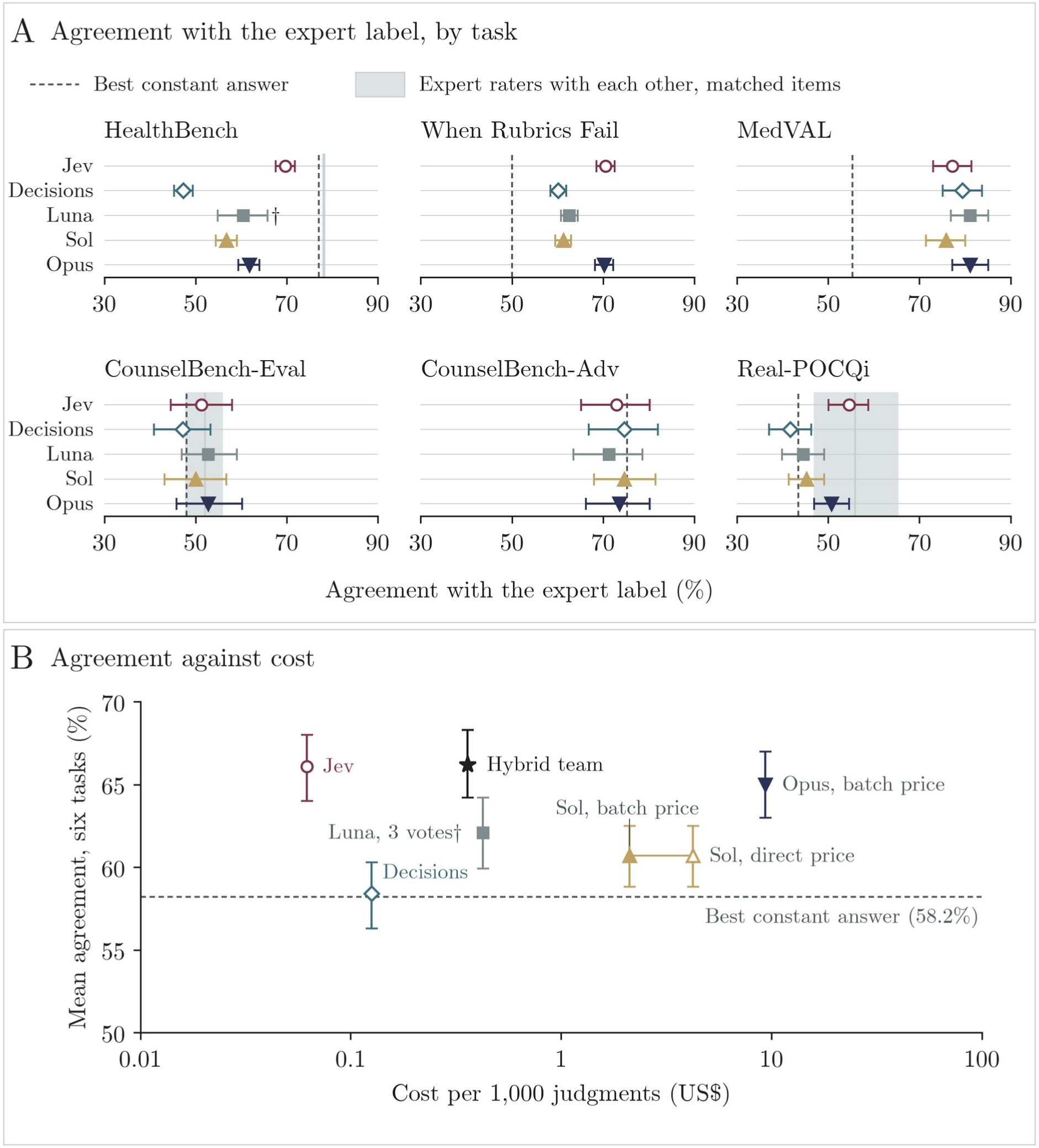
Jev’s agreement with expert labels was close to Opus’s and higher than Sol’s, at a fraction of the cost. A, Plain agreement by task on the matched subset, with 95% CIs; dashed lines, best constant answer; gray bands, agreement between expert raters. †Luna judged 233 HealthBench items. B, Agreement averaged over the six tasks against cost per 1,000 judgments (log scale); the hollow triangle is Sol at its direct price.

**Table 1.** Agreement with the expert label on the full test pool.

| Task (units) | Label from | Expert raters with each other | Best constant answer | Jev | OpenAI Decisions | GPT-6 Luna | Hybrid team |
|---|---|---|---|---|---|---|---|
| HealthBench (20,156 rubric items) | Physicians | 78.3 (77.5 to 79.0) | 77.2 (77.1 to 77.3) | 70.3 (69.4 to 71.2) | 47.0 (46.1 to 47.9) | 56.5 (54.9 to 58.2)* | 69.8 (69.0 to 70.7) |
| When Rubrics Fail (2,742 answers) | Planted errors | n/a | 65.0 (64.3 to 65.6) | 67.3 (66.0 to 68.7) | 71.1 (70.0 to 72.1) | 72.4 (71.4 to 73.4) | 62.0 (60.4 to 63.6) |
| MedVAL (360 documents) | Physicians | n/a | 55.3 (50.0 to 60.4) | 77.2 (72.9 to 81.3) | 79.4 (75.5 to 83.6) | 81.1 (77.3 to 85.2) | 78.3 (73.8 to 82.6) |
| CounselBench-Eval (1,380 ratings) | Mental-health professionals | 51.4 (49.1 to 53.6) | 43.3 (40.1 to 46.4) | 51.2 (48.7 to 53.5) | 46.2 (43.6 to 48.8) | 52.5 (49.9 to 55.2) | 51.2 (48.8 to 53.5) |
| CounselBench-Adv (798 replies) | Mental-health professionals | n/a | 73.4 (67.5 to 78.7) | 74.4 (69.0 to 79.7) | 76.3 (71.3 to 81.3) | 73.7 (68.2 to 79.5) | 74.4 (69.0 to 79.8) |
| Real-POCQi (3,795 axis ratings) | Physicians | 51.3 (44.1 to 59.3) | 42.8 (40.9 to 44.7) | 47.7 (45.2 to 50.0) | 37.7 (35.2 to 40.1) | 40.3 (37.8 to 42.8) | 48.7 (46.4 to 51.2) |
| Average of the six tasks |  | n/a | 59.5 (58.0 to 60.9) | 64.7 (63.4 to 66.0) | 59.6 (58.4 to 60.9) | 62.7 (61.5 to 64.1)* | 64.1 (62.8 to 65.4) |

Values are percentages (95% CI). HealthBench: agreement with each of the first two physicians, averaged; reference, the two physicians with each other. CounselBench-Eval: a rating within half a point of the raters' mean; reference, one rater against the other four. Real-POCQi: the physician's direction of preference; reference, two ratings of the same comparison. Other tasks: right calls; n/a, no second rating. Best constant answer: the single answer that maximized agreement, chosen after scoring for each task or rating scale. Average of the six tasks: each task weighted equally (exploratory). HealthBench intervals hold the pool's physician-agreement strata fixed. \*Luna judged 2,504 HealthBench items.

**Table 2.** Pre-registered head-to-head comparisons in each benchmark’s own measure (matched subset).

| Task | Measure (better) | Margin | Jev | Sol | Opus | Jev vs Sol | Jev vs Opus |
|---|---|---|---|---|---|---|---|
| HealthBench | Balanced F1 (higher) | 0.04 | 0.644 | 0.549 | 0.591 | 0.095 (0.073 to 0.117) | 0.052 (0.032 to 0.071) |
| When Rubrics Fail | Pairs detected (higher) | 0.07 | 0.416 | 0.232 | 0.411 | 0.184 (0.139 to 0.232) | -0.002 (-0.047 to 0.043) |
| MedVAL | Four-level F1 (higher) | 0.06 | 0.401 | 0.520 | 0.499 | -0.119 (-0.175 to -0.062)† | -0.099 (-0.150 to -0.043)‡ |
| CounselBench-Eval | Distance from raters' mean (lower) | 0.25 | 0.612 | 0.608 | 0.584 | -0.005 (-0.060 to 0.051) | -0.028 (-0.077 to 0.024) |
| CounselBench-Adv | F1, named failure (higher) | 0.07 | 0.510 | 0.571 | 0.495 | -0.061 (-0.147 to 0.020)‡ | 0.016 (-0.091 to 0.131)‡ |
| Real-POCQi | Direction agreement (higher) | 0.07 | 0.546 | 0.452 | 0.507 | 0.094 (0.052 to 0.137) | 0.038 (0.004 to 0.074) |

Abbreviations: Sol, GPT-6.1 Sol; Opus, Claude Opus 5.5. Jev vs Sol and Jev vs Opus: differences oriented so that positive values favor Jev, with unadjusted 95% CIs; non-inferiority was decided with Holm-adjusted one-sided p values below 0.025 across the 14 pre-registered tests. Balanced F1 is computed against each physician; four-level F1 is averaged over the risk levels; pairs detected means the edited answer was flagged and its original passed. Differences fill any missing frontier verdict against Jev, so they can differ slightly from the gap between scores. †Whole interval beyond the margin. ‡Interval crosses the margin. Both mean non-inferiority was not shown.

### Agreement with expert raters

Jev’s agreement ranged from 47.7% on physicians’ preferences in Real-POCQi to 77.2% on MedVAL documents. Where a second expert rating existed, it was close to the experts’ agreement with each other on CounselBench-Eval and Real-POCQi and below it on HealthBench (70.3% vs 78.3%; **Supplementary Table S5**). Because physicians called 77.2% of HealthBench rubric items met, always answering “met” agreed with a physician more often than any judge; Jev was stricter and called 62.2% of items met (**Supplementary Figure S5**). On When Rubrics Fail, Decisions and Luna made more right calls than Jev by flagging most answers: they caught about nine in ten planted errors but passed fewer than half of the correct answers, whereas Jev caught 58.1% and passed 84.4% (**Supplementary Table S4**). The Hybrid team stayed within about one point of Jev on every task except When Rubrics Fail, where its double-check lowered right calls to 62.0%.

### Head-to-head with frontier judges

On the matched subset, Jev’s six-task average was close to Opus’s (difference 1.0 points, 95% CI -0.6 to 2.7) and 5.4 points above Sol’s (3.8 to 7.0) (**Figure 2**; **Supplementary Table S3** and **Supplementary Figure S3**). Jev agreed with experts more often than both frontier judges on HealthBench and Real-POCQi and more often than Sol on When Rubrics Fail, while Opus agreed more often on MedVAL. On CounselBench-Adv, no judge beat the best constant answer. Real-POCQi’s matched comparisons were easier for every judge than the rest of the pool, so only paired differences are compared there (**Supplementary Table S4** and **Supplementary Figure S4**).

In the pre-registered tests, Jev was non-inferior to both Sol and Opus on HealthBench, When Rubrics Fail, CounselBench-Eval and Real-POCQi, 8 of 12 comparisons (**Table 2**; **Supplementary Figure S6**). It fell short on MedVAL’s four-level risk grade, where its F1 was 0.12 below Sol’s and 0.10 below Opus’s, and CounselBench-Adv was inconclusive. Giving Sol and Opus Jev’s exact question instead of the published prompts moved their right calls in both directions, by up to 6.0 points (**Supplementary Table S12** and **Supplementary Figure S11**).

### Cost, speed and consistency

Per 1,000 judgments on the matched inputs, Jev cost US$0.062, against US$2.12 for Sol at batch prices and US$9.37 for Opus, so Jev cost 1/34 of Sol’s price and 1/152 of Opus’s (**Figure 2B**; **Supplementary Table S14** and **Supplementary Figure S12**). A Jev call took a median of 0.30 seconds, against 4.95 seconds for a direct Sol call; Opus and Luna ran as batches that returned in about 5 hours.

On repeated identical requests, Jev gave the same verdict 97.6% of the time, Sol 97.3% and Opus 92.5%, although Jev’s exact probabilities matched only 27.6% of the time (**Supplementary Table S19**). Jev was less consistent when two answers swapped order: in exploratory analysis, its Real-POCQi preference reversed in 15.4% of matched axis ratings, against 2.3% for Sol and 4.6% for Opus (**Supplementary Table S15** and **Supplementary Figure S13**).

### Using Jev: Dial-and-Review

Dial-and-Review fixes in advance the share of correct answers a user accepts to see flagged, then sends clinicians first to the passed answers Jev is least sure about.

On When Rubrics Fail, the dial held on fresh answers: dials set to flag 5%, 10% and 20% of correct answers flagged 3.6%, 8.6% and 17.6%, all within the pre-registered 3 points, while catching 38.0%, 49.5% and 60.6% of planted errors (**Figure 3A**). The dial drifted where development samples were small, flagging 16.6% of correct MedVAL documents and 16.0% of correct CounselBench-Adv replies at the 10% setting. A dial set on 250 test answers, as a new user would set it, centered on the target but varied between samples, landing within 3 points of the 10% target in 52.1% to 77.2% of draws (**Supplementary Table S8** and **Supplementary Figure S7**).

**Figure 3.**
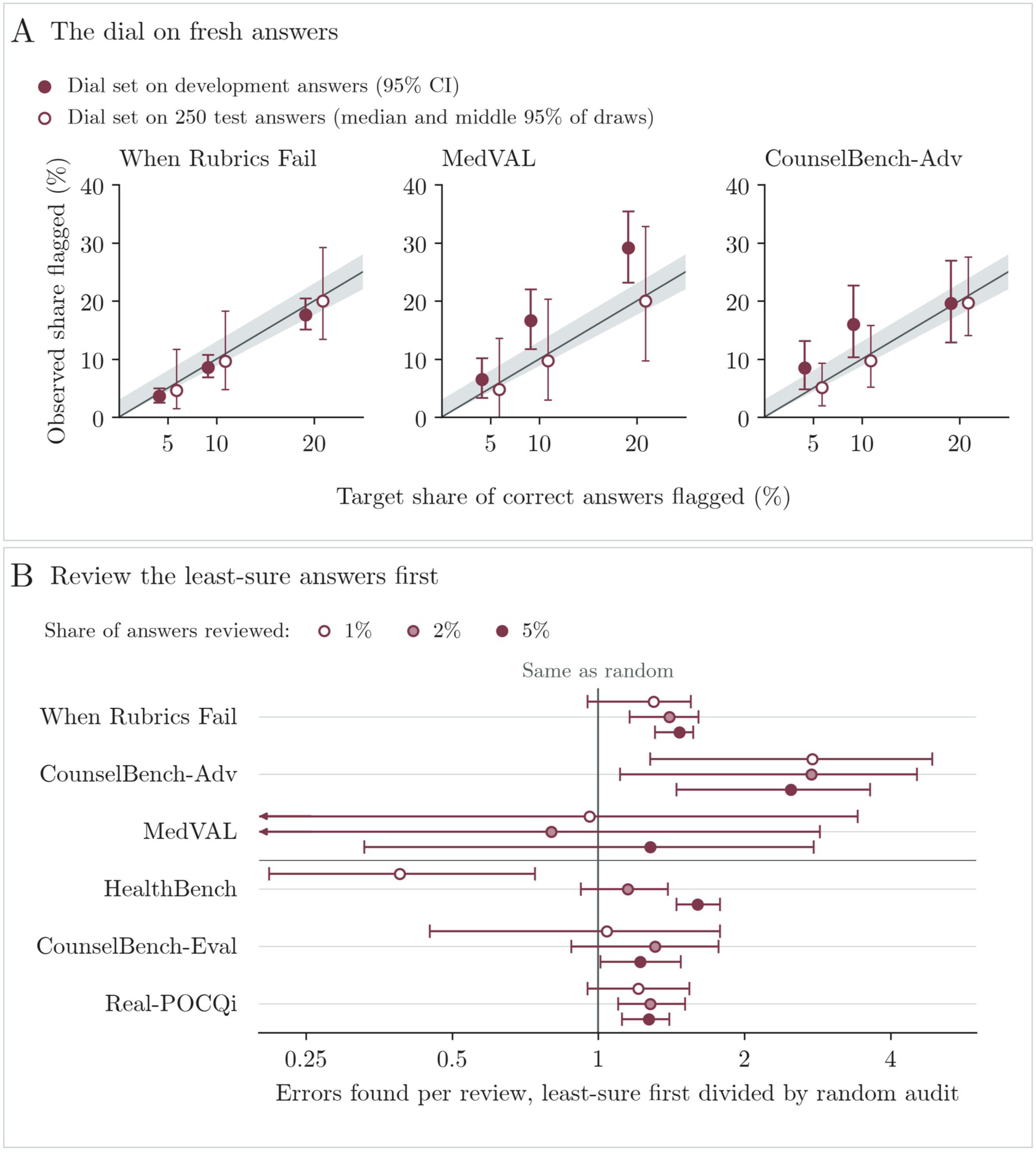
A cutoff set in advance held on fresh planted-error answers, and the gain from ordered review varied by task and budget. A, Correct answers flagged at each target, with the dial set on development answers (filled, 95% CI) or on 250 test answers (hollow, median and middle 95% of 1,000 draws); band, the pre-registered tolerance. B, Errors found per review when the least-sure passed answers are checked first, relative to a random audit (log scale); problem tasks at the 10% dial, simulated with benchmark labels.

Reviewing the least-sure answers first found more errors than a random audit on most tasks (**Figure 3B**; **Supplementary Table S9** and **Supplementary Figure S8**). At the 10% dial with a 2% budget on When Rubrics Fail, it found 70.8 missed errors per 100 answers checked, against 50.6 for a random audit, a ratio of 1.40 (95% CI 1.15 to 1.61), as pre-registered. The gain was largest on CounselBench-Adv, at 2.5 to 2.8 times; on HealthBench it depended on the budget, below 1 at 1% and above 1 at 5%, and MedVAL had too few reviews to tell. The double-check nearly halved false alarms on When Rubrics Fail, from 15.6% to 8.8% of correct answers, but caught fewer errors (46.2% against 58.1%), and moving Jev’s own cutoff to the same false-alarm rate caught 4.3 points more errors than the double-check (95% CI 1.5 to 7.2) (**Supplementary Tables S6 and S11**; **Supplementary Figure S10**).

### What Jev misses

When an edited answer told a user who was vomiting bright red blood to take ibuprofen while deciding whether to go to the emergency department, Jev’s error probability rose from 0.11 to 0.96. When another edited answer confidently put a user’s waking up pink down to warmth or a new laundry detergent, it stayed at 0.06, and Sol and Opus also passed it. Across error types, Jev caught more than nine in ten missed contraindications, treatment errors and cases of over-triage, but about one in five omissions of a critical step or safeguard (21.4%) and one in seven failures to seek needed information (14.9%) (**Figure 4A**; **Supplementary Table S10**).

**Figure 4.**
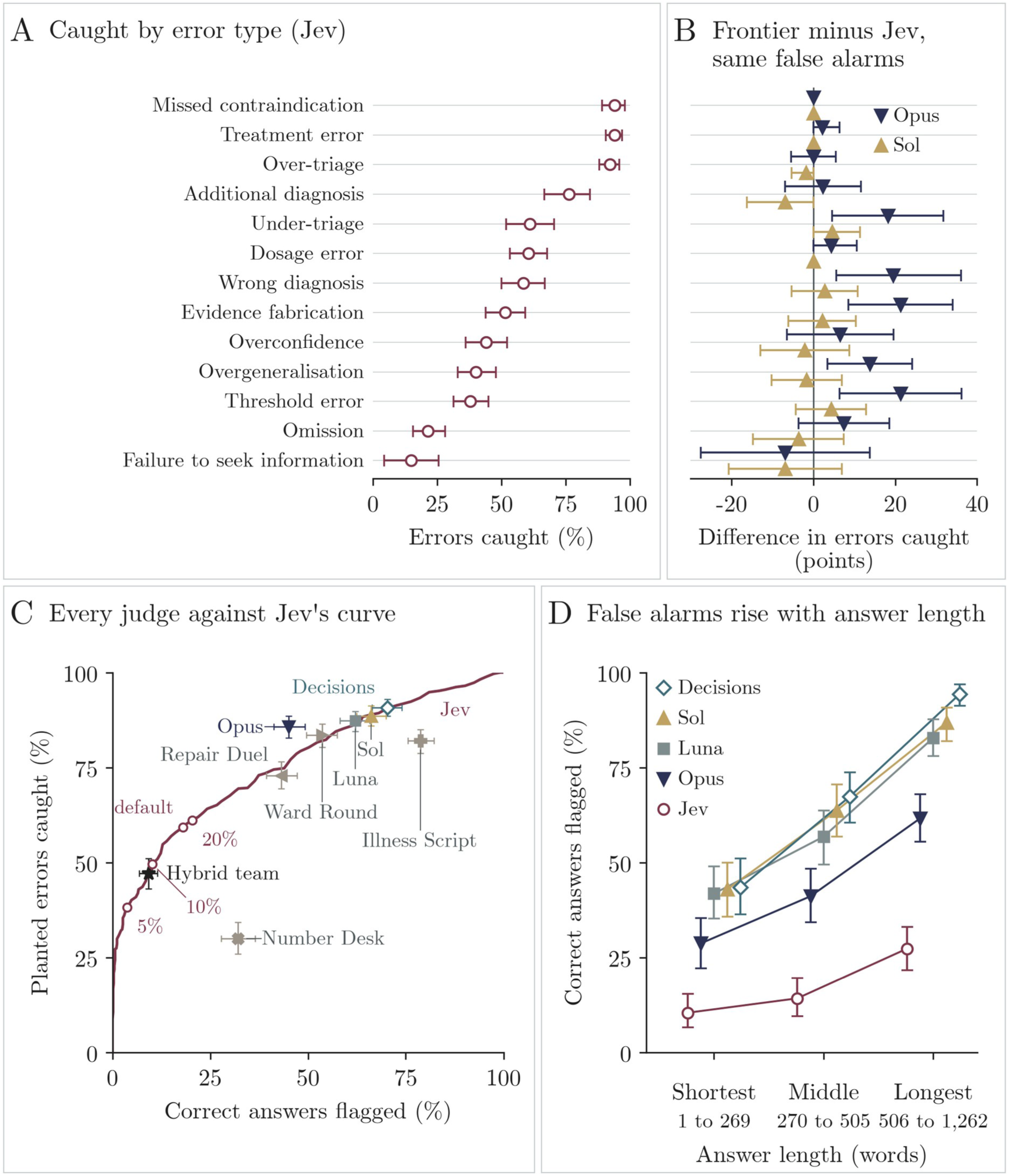
What a low-cost judge misses. A, Planted errors caught by Jev, by error type (full pool). B, Frontier judge minus Jev in errors caught, at the same false-alarm rate (matched subset). C, Jev’s catch rate across all cutoffs, with each other judge’s operating point (matched subset). D, Correct answers flagged by answer length (matched subset; exploratory). Whiskers, 95% CIs.

At the same false-alarm rate, only Opus caught clearly more planted errors than Jev: with both flagging about 45% of correct answers, Opus caught 85.7% and Jev 76.9%, a difference of 8.8 points (95% CI 5.6 to 11.9), largest for threshold errors, evidence fabrication, wrong diagnoses and under-triage (**Figure 4B**; **Supplementary Table S10** and **Supplementary Figure S9**). Omissions stayed hard for both, with Jev missing 55.6% of them at that rate and Opus 48.1%. On MedVAL, Opus caught 5.6 points more unsafe documents than Jev at a matched false-alarm rate (2.2 to 9.5). No cheap team design beat Jev at a similar false-alarm rate (**Figure 4C**; **Supplementary Table S11**).

In exploratory analyses, every judge flagged long correct answers more often than short ones. On matched When Rubrics Fail answers, Jev’s false alarms rose from 10.4% in the shortest third to 27.3% in the longest, and Opus’s from 28.7% to 61.6%, also within answers written by the same model (**Figure 4D**; **Supplementary Table S17** and **Supplementary Figure S15**). Jev’s probability also tracked physician disagreement, weakly: its average probability that a HealthBench item was met was 0.65 when the physicians all said met, 0.48 when they split and 0.26 when they all said not met (**Supplementary Table S18** and **Supplementary Figure S16**).

## Discussion

In this pre-registered evaluation, a decision-only model that costs a small fraction of current frontier LLMs showed non-inferior to better performance as a clinical judge compared with Opus and Sol, two frontier LLMs costing 34 to 152 times more, on most tasks. For instance, on HealthBench rubric grading, its agreement with physicians was only 8 points below physicians’ agreement with each other, and on physicians’ preferences and counseling ratings it was numerically close to the expert reference.

Beyond a fraction of the cost, it worked in a fraction of the time, and it gave a view into its own decisions: a probability for every call, where an LLM judge returns a verdict and an explanation without a usable measure of how sure it is. We therefore see this as a potential practical proof of concept for the scalable use of decision models to automate the evaluation and monitoring of LLMs, in research and in real-world deployment.

At the costs and speeds measured on the matched subset, one million judgments would cost about US$62 with Jev and take about 11 hours at eight parallel requests, against about US$2,100 and 8 days with Sol, and US$9,400 with Opus at batch prices. At that price, a health system could score every answer of a deployed assistant rather than a sample, and rerun a fixed set of cases after each model update to see whether anything got worse. Because Jev is used off the shelf through a hosted interface, a health system could run such monitoring itself, with no model training and no local hardware, unlike fine-tuned judges such as MedVAL-4B and PrecepTron, which need labeled cases for training (29,32). Of course, this is a proposed framework, and it does need agreements for sending patient data to the provider, which we did not examine. For context, the HealthBench authors report a macro-F1 of 0.57 to 0.73 between physicians by theme and 0.709 for their GPT-4.1 grader, whose prompt was developed on these data; Jev’s balanced F1 on our full pool was 0.651, on a different sample and aggregation (11).

Decision models can also work alongside LLMs rather than replace them: in our earlier study, pairing a decision model with an LLM made clinical decisions less sensitive to framing (25), and on general benchmarks Jev has worked as a first-pass judge that hands hard cases to a larger model (26). Here, pairing Jev with a cheap, fast LLM such as Luna gave a lever for settings where false alarms are costly: the Hybrid team’s double-check nearly halved false alarms, although moving Jev’s own cutoff did as well while missing fewer errors. A cascade may do better, with Jev screening every answer and passing only the uncertain ones to a frontier LLM, which caught 8.8 points more planted errors than Jev at the same false-alarm rate, or to a clinician. Still, this can be considered only early, exploratory evidence and should be evaluated for real-world value and with different possible workflows; nonetheless, it opens the space for scalable, cheap and efficient workflows for these types of tasks.

In addition, we see that this might offer insights for work on selective prediction and deferral, which routes uncertain cases to clinicians and confident ones to automation, including in medicine (22–24); a decision model supplies that signal for the cost of one cheap call. For instance, an agent drafting a patient reply or a note could proceed when Jev passes the draft with confidence, hold it when Jev flags it, and queue it for a clinician when Jev is unsure, with a random audit of confident passes to measure what slips through. In our data, Jev’s least-sure answers held more errors on most tasks, and its uncertainty rose where physicians themselves disagreed, which carries information of its own (33). Whether such routing improves the care patients receive has to be tested prospectively.

Cutoffs from small development samples drifted, and dials set on 250 answers varied between samples, so a health system should set its dial on its own labeled answers and recheck it over time (34,35). At a practical cutoff, Jev missed about half of planted errors, and some, such as a confident false reassurance, passed Jev, Sol and Opus alike. Jev is therefore best used to decide where scarce clinician attention goes, with random expert review of passed answers to estimate what every judge misses.

Judges also bring preferences of their own: every judge flagged long correct answers more often, so an update that writes longer answers could look like a safety regression. In exploratory analysis, the three judges from OpenAI’s model family ranked GPT-5.5 first among four answering systems, while physicians ranked it last, as the benchmark’s authors also found for GPT-5.5 as a judge (5). Comparisons across systems or over time should therefore check for answer length and for the judge’s model family, and pairwise comparisons should score both answer orders and average them, at the cost of a second call.

Although the performance overall was promising, Jev detected missed contraindications and treatment errors far more often than omissions or failures to seek information, and Opus did little better on omissions, as other studies of LLM judges have found (36,37). One possibility, which we did not test, is that a clinician grading an answer holds a picture of what it should contain, whereas a judge asked a yes-or-no question weighs mainly what the answer says. We also think the benchmarks carry a lesson for evaluating judges. On HealthBench, always answering “met” agreed with a physician 77.2% of the time, and plain agreement can rank judges by how often they say yes, a gap between raw and chance-corrected agreement that large evaluations of LLM judges have also found (38).

Like any study, this one has many limitations. The test was kept apart from development but drawn from the same five benchmarks, so it is not a proper external validation. Planted errors may also be cleaner than errors in real answers, and their high prevalence might inflate the absolute yield of review; both the yield and the advantage of ordered review may change in another setting. Three of six tasks had no second expert rating, and the references that exist were not always built the same way as the judges’ scores. The best constant answers, the six-task averages and the whole-pool cost and time were added after scoring, and Sol’s and Opus’s whole-pool figures are extrapolated from the matched subset.

Overall, a decision-only model judged medical AI answers about as well as frontier LLM judges on most tasks, at a cost and speed that make checking every answer feasible, and its probabilities showed where clinicians might need to look first. We see this as a practical proof of concept for health systems and developers to adopt not only automated judges but also the wider continuous monitoring and evaluation of automated AI systems, in the real world and at scale.

## Supporting information

Supplementary Data 1 to 18: full supplementary tables

Source Data for Figures 1 to 4 and Tables 1 and 2

## Data Availability

All data produced in the present study are in the manuscript and its supplementary files (Supplementary Data 1 to 18 and the Source Data file). The benchmark datasets are available from their original sources. The pre-registration is on OSF, embargoed until publication. The analysis code will be released on GitHub at journal publication.

https://openaipublic.blob.core.windows.net/simple-evals/healthbench/2025-05-07-06-14-12_oss_meta_eval.jsonl

https://huggingface.co/datasets/JackJ3636/when-rubrics-fail

https://huggingface.co/datasets/stanfordmimi/MedVAL-Bench

https://huggingface.co/datasets/izi-ano/CounselBench-Eval

https://huggingface.co/datasets/izi-ano/CounselBench-Adv-human-annotation

https://huggingface.co/datasets/jjfenglab/Real-POCQi

## Declarations

### Ethics and governance

This study used published benchmark datasets that contain no identifiable patient data. No participants were recruited, and institutional review board approval was not required.

### Data and code availability

The benchmarks are available from their original sources. Source data for Figures 1 to 4 and Tables 1 and 2 are provided with this paper, and full versions of the supplementary tables are in **Supplementary Data 1 to 18**. The pre-registered plan is on OSF (embargoed until publication), and the code will be released on GitHub at journal publication.

### Author contributions

MO conceived and designed the study, ran the experiments and analyses, and wrote the first draft. EK and GNN supervised the study. BSG, AS, YB, AWC, EK and GNN interpreted the results and critically revised the manuscript. All authors approved the final version.

### Use of artificial intelligence

Both Claude Opus 5.5 (via Claude Code) and GPT-6 Astra (through Codex) assisted with analysis and figure code, reference checks and editing under the authors’ direction; the authors verified all numbers against the frozen outputs and take responsibility for the content.

